# The RESPONDER trial: A feasibility study of an early intervention model deploying community first responders to administer intranasal naloxone in suspected opioid overdoses

**DOI:** 10.64898/2025.12.19.25342601

**Authors:** Julia Rehn, Cecilia Andréll, Josephine Hansson, Katja Troberg, Pernilla Isendahl, Martin Bråbäck, Leif Svensson, Fredrik Byrsell, Andreas Claesson, Anders Håkansson

**Affiliations:** Malmö Addiction Centre, Skåne University Hospital, Region Skåne, Södra Förstadsgatan 35, 205 02 Malmö, Sweden; Department of Clinical Sciences Lund, Psychiatry, Faculty of Medicine, Lund University, Sölvegatan 19, 221 84 Lund, Sweden; Centre for Cardiac Arrest, Anesthesiology and Intensive Care, Department of Clinical Sciences Lund, Faculty of Medicine, Lund University, Sölvegatan 19, 221 84 Lund, Sweden; Practicum Clinical Skills Centre, Team CPR, Skåne University Hospital, Malmö, Sweden; Department of Infectious Diseases, Skåne University Hospital, Malmö, Sweden; Centre for Resuscitation Science, Department of Clinical Science and Education, Södersjukhuset, Karolinska Institutet, Stockholm, Sweden; SOS Alarm AB, Stockholm, Sweden

**Keywords:** Opioid overdose, community first responder, volunteer first responder, emergency medical services, emergency medical dispatch, *Drug Overdose/diagnosis/drug therapy/epidemiology, *Naloxone/therapeutic use, smartphone alerting system, first responder system

## Abstract

**Introduction:** Opioid use continues to cause harm and fatalities worldwide, despite ongoing preventive efforts. Early administration of the antidote naloxone can reverse potentially fatal opioid overdoses. Naloxone distribution programs have increased survival across regions; however, additional efforts are needed to reduce opioid overdose mortality. One approach is to expand naloxone availability within communities.

**Methods and analysis:** The feasibility trial RESPONDER (REgion Skåne Preventing Overdose through Naloxone Distribution with Emergency Runners) introduces a unique smartphone alerting system with community first responders (CFRs) dispatched to suspected opioid overdoses, in addition to dispatch of regular emergency medical services (EMS). The aim is to investigate if trained CFRs can successfully recognize and reverse overdoses caused by opioids, prior to EMS arrival. The CFRs will be equipped with nasal naloxone and introduced to a novel naloxone algorithm during a study-specific course prior to participation. The course will offer practical training in basic life support and first aid, and it will also assess competence in the low-arousal approach, as well as the legal and ethical aspects of being a CFR.

Main outcomes of the trial are feasibility, acceptability and safety. The trial will be performed in the region of Skåne, Sweden, between September 1st, 2025, and August 31st, 2027, with every CFR followed for up to 12 months.

**Ethics and dissemination:** The trial has been approved by The Swedish Ethical Review Authority (file number 2024-05887-01). Written informed consent is required from participating CFRs. This is also required of overdose survivors to allow the collection of clinical data from hospital records.

Information regarding the project and recruitment will be disseminated via social media, news media, through the healthcare region’s communication channels, and within public transportation hubs. Outcomes and analyses will be submitted to peer-reviewed journals.

**Conclusion:** Safe, early and effective management of opioid overdoses by trained CFRs prior to EMS arrival may reduce morbidity and mortality among people who use opioids. Findings from the current trial may contribute to further knowledge on how to implement and improve CFR systems for opioid overdoses globally. ClinicalTrials.gov-ID: NCT07079241.

**Article Summary:** *Strengths and limitations of this study:* - The RESPONDER trial introduces a novel community first responder system for time-critical opioid overdose reversal in addition to out-of-hospital cardiac arrest. - The trial aims to enhance access to naloxone and raise community awareness, improving the likelihood of early and potentially life-saving overdose reversal. - A study-specific course introduces a novel naloxone algorithm which has been developed by expertise in the fields of resuscitation, First Aid and opioid use disorder. - The study group consists of researchers and clinicians with a broad range of expertise concerning opioid overdose management, substance use disorders, First Aid education, emergency medical services, emergency medical dispatch centre operations and community first responder systems, adding several in-depth perspectives to this community first responder model. - A limitation may be that overdose events unreported to emergency dispatch centres will not reach the trial’s community first responders.

## INTRODUCTION

### Opioid overdoses: morbidity and mortality

Opioid use disorder represents a major public health concern, adversely impacting quality of life for individuals worldwide (1). The situation in the United States has been referred to as an opioid epidemic, while high rates of opioid-related morbidity and mortality have also been reported in other countries. In 2023, the European Monitoring Centre for Drugs and Drug Addiction reported toxicological findings of opioids post-mortem in 7 out of 10 analyzed fatal overdoses in the European Union (2). The Public Health Agency of Sweden reported an estimation of 474 narcotics-related fatalities in Sweden 2023, among which 82% of the toxicologic analyses involved opioids, and the mortality rate for opioid overdose in Sweden are above the average rate in Europe (2–4). Polydrug use, especially opioids combined with sedatives, contributes to the high mortality rates through synergistic respiratory inhibition. Between 36 and 74% of people with opioid use disorder have themselves experienced a non-fatal overdose (5–9), while over 80% have witnessed at least one opioid overdose (6, 7). Darke et al. estimated an incidence of 20-30 non-fatal opioid overdoses among heroin users for every single fatal overdose (10). International overdose mortality rates are three to four times higher among men than women (2). In Sweden, gender distribution is comparable, and both the mean and median age at death are approximately 40 years (3, 11). Different surveillance methods are used throughout the research field and the total overdose incidence, non-fatal and fatal, is consequently suspected to be highly underestimated (12–15).

Non-fatal opioid overdoses frequently lead to serious complications, including hypoxia-induced brain damage. (16, 17). Opioid-related respiratory arrest can progress within minutes, making timely overdose reversal critical (14, 17). Untreated respiratory arrest may culminate in cardiac arrest (18, 19). The Swedish Cardiac Arrest Registry reported 125 out-of-hospital cardiac arrests (OHCA) due to intoxication, involving opioid overdose, in 2024, and 176 cases in 2023 (20).

### Effective interventions to reduce opioid overdose-related morbidity and mortality

The competitive antagonist naloxone counteracts the respiratory depression caused by opioids. Naloxone has been proven safe to use in various administration formats and doses, also in pediatric populations (21, 22). Adverse events of naloxone are mostly related to acute withdrawal from long-term opioid use (17, 23). Opioid withdrawal can be profoundly distressing and painful to experience. Withdrawal is characterized by reduced concentration and stress tolerance, increased emotional lability, and physical symptoms such as sweating, rhinorrhea, nausea, diarrhea and chills (17, 24). Symptomatic treatment can partially alleviate withdrawal symptoms. Withdrawal typically lasts for several days in the absence of opioid consumption. The symptoms increase in a dose-dependent manner during administration of naloxone among individuals with long-term opioid use, until the naloxone has bound to all opioid receptors.

However, naloxone is typically eliminated more rapidly than opioids (17, 21). Repeated administration of naloxone may be required, highlighting the need for prolonged observation, for example, in a hospital setting following opioid overdose.

Naloxone can be administered via several routes: intravenous, subcutaneous, intramuscular injections, and intranasal. Intranasal naloxone spray has simplified administration by non-medical professionals (25). The concept of take-home naloxone (THN) has revolutionized the field, with naloxone distribution programs worldwide showing promising results for safety and feasibility (26–28). Large-scale follow-up studies are few, but strong political support for THN exists (23, 26, 29, 30). The United Nations has declared distribution of naloxone to be an evidence-based strategy for opioid overdose reversals (31).

THN programs were introduced in Sweden in 2018, followed by national guidelines in 2019 that promoted the distribution of naloxone to individuals at risk of experiencing opioid overdoses (32–34). Internationally, the American Heart Association released updated guidelines in 2025 recommending increased public access to naloxone (35).

A regional naloxone program in Skåne, Sweden, distributed around 18,600 doses of intranasal naloxone to more than 3,360 individuals between June 2018 and June 2025. Over 1,500 overdose self-reported reversals have been reported (Personal communication, June 26th, 2025: Isendahl, P.). A decreasing trend in fatal overdoses was observed, but it was significant only among men. (11). Nasal naloxone was from March 2024 available for adults to purchase without prescription at Swedish pharmacies (36). Prior international reports have addressed challenges with prescription-free naloxone in terms of cost and stigma (37, 38). The implementation in Sweden is yet to be evaluated.

### Emergency dispatch systems with community first responders

Although the THN concept is well-established in many countries, coverage is insufficient to reduce mortality and morbidity of opioid overdoses. Naloxone is currently recommended to be distributed to people at risk of experiencing an overdose in Sweden (36). This contrasts to international guidelines, which recommends naloxone to be distributed to both people who use drugs, and to their family and friends who are at risk of witnessing an overdose (23). It has been suggested that lay persons without any prior relationship to people at risk of experiencing an overdose should have the opportunity to carry naloxone as well (35, 39). The concept is based on successful dispatch of community first responders (CFRs), using a smartphone application, to suspected OHCAs. The terminology of CFR has varied in the prior literature. The current paper will use the suggested definition *community first responder*, following an international consensus study by Metelmann, C. et al., 2025 (40).

Prior CFR dispatch systems for OHCA have demonstrated improved survival, fewer complications, and a significant time advantage for initiating cardiopulmonary resuscitation (CPR) and automated external defibrillator (AED) use before emergency medical services (EMS) arrival (41–43). Qualitative research with CFRs indicates a strong willingness to assist in emergencies, while adverse psychological effects at the individual level appear to be rare (44, 45). Only in Sweden, 2025, a smartphone-based CFR service for OHCAs has 150,000 registered CFRs, including currently around 24,000 users in Region Skåne. At least one CFR accepts the OHCA dispatch in 98% of all cases in Skåne (Personal communication with the operations manager in Skåne, November 11th, 2025, Andréll, C.).

CFR services are found in several parts of Europe, as well as in Asia and North America (46). The RESPONDER (REgion Skåne Preventing Overdose through Naloxone Distribution with Emergency Runners) trial extends the CFR concept to include assistance for opioid overdoses alongside OHCAs (Figure 1).

**Figure 1.**
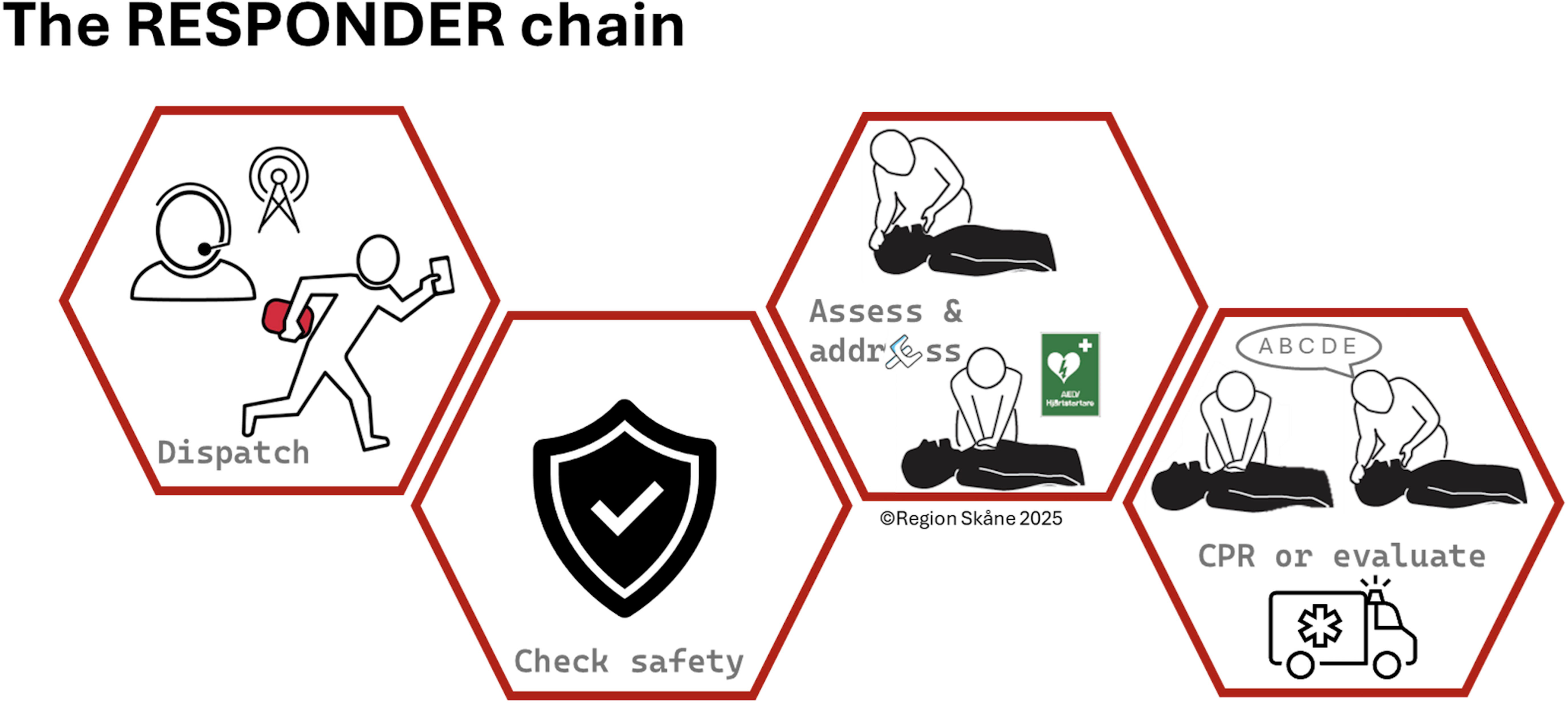
The RESPONDER chain (1) Trained community first responders (CFR) with naloxone are dispatched by the Swedish emergency medical dispatch service to suspected opioid overdoses, in addition to out-of-hospital cardiac arrests, (2-4) CFR arrives and follows the study-specific naloxone algorithm

### Objectives

#### Aims & research questions

Our primary aim is to assess the feasibility, acceptability and safety of a smartphone application connected to the Swedish emergency dispatch service and dispatching trained CFRs to suspected opioid overdoses. The main research question is: Is it feasible and safe to dispatch First Aid-trained CFRs with in-depth opioid and overdose knowledge, including legal and ethical aspects, equipped with nasal naloxone, to the scene of suspected opioid overdoses before EMS arrival?

### Hypotheses

#### We hypothesize that a novel CFR dispatch system for opioid overdoses is

- Feasible with sufficient recruitment, technological positioning and dispatching of First Aid- and overdose-educated CFRs equipped with naloxone
- Time-effective with overdose reversal by CFRs before EMS arrival
- Accepted by CFRs and overdose victims as a beneficial method to reverse overdoses
- Safe with low degree of adverse events and high acceptability among both CFRs and overdose victims

#### Trial design

The RESPONDER trial is an interventional feasibility study of a two-year pilot project. At least five different sub-studies will be performed during and 1-2 years after the pilot period. The quantitative studies will primarily explore feasibility, acceptability and safety of the intervention. Two qualitative studies will examine: (1) overdose survivors’ experiences after receiving naloxone from a CFR, and (2) CFRs’ experiences of accepting overdose-related dispatches. The two viewpoints will be addressed in separate studies.

## MATERIALS AND METHODS

### Study setting

The RESPONDER trial will be conducted in the region of Skåne, Sweden, between September 1, 2025, and August 31, 2027. Skåne has around 1.43 million residents, and one fourth of them live in the biggest city, Malmö.

### Emergency medical dispatch in Sweden

Emergency 112-calls in Sweden’s 21 regions are answered by trained emergency dispatchers at overall 14 emergency medical dispatch centres (EMDCs) throughout the country (47). The dispatchers answering emergency calls may be located at any of the EMDCs, as calls are routed through a national queueing system. The call is interconnected at an early stage to a regional ambulance dispatcher if a priority 1A- or 1B-level condition is recognized, thus resulting in the immediate dispatch of EMS-units to the scene. Most regions in Sweden, including Skåne, follow a one-step procedure in which the dispatcher manages the emergency call from start to finish. If needed, the dispatcher can consult a registered nurse at the EMDC to assist in determining the appropriate triage level (47).

Dispatch codes and procedures, including telephone-guided instructions to the calling bystander, are supported by a national decision-making tool implemented 2021-2022 called STEP (48).

Dispatchers at EMDCs using the one-step procedure in Sweden must complete a 13-week training program; however, prior medical employment experience is not required (47).

### The RESPONDER system

The intervention of the RESPONDER trial is based upon a smartphone alerting system with CFRs manually dispatched to suspected opioid overdoses and OHCAs, as a complement to standard of care with dispatch of EMS.

The CFR system for suspected opioid overdoses used in this trial is developed from the existing system by Heartrunner Sweden AB (49). The existing system dispatches CFRs to suspected OHCAs in Denmark’s five regions and to 14 out of 21 regions in Sweden.

The functionality of the RESPONDER system is illustrated in Figure 1. An emergency call to the EMDC initiates a chain of events. While EMS is being dispatched to the scene, the application’s positioning system identifies up to 30 CFRs within a radius of 10,000 meters from a suspected overdose, or within 1,500 meters from a suspected OHCA. These radii may be adjusted if an insufficient or an excessive number of CFRs arrive before EMS. The current Swedish application, which is used exclusively for OHCAs, dispatches CFRs at the same time as the RESPONDER application. This explains the initially shorter radius from OHCAs in our study.

The application provides information on the nearest route to the scene, together with information on distance and accessible AEDs in the vicinity.

People who volunteer to become CFRs in the trial will attend a one-day course, including the following:

### Study-specific part of the course, mandatory for all participants

- Presentation of opioid characteristics and importance of overdose reversal including statistics and ongoing efforts to reduce opioid-related morbidity and mortality
- Recognition and management of opioid overdoses through administration of naloxone and initiating CPR, including technique to handle mouth-to-mask ventilation as well as compliance to and administration of naloxone both safe and correct
- Practical training in administration of naloxone and CPR, following a novel study-specific naloxone algorithm (Figure 2) which was inspired by a recently updated First Aid guideline from American Heart Association (50) and further developed by expertise in the fields of resuscitation, First Aid and opioid use disorder
- Ethical and legal considerations in emergency settings
- Strategies to safely approach emergencies and people in distress, following a low-arousal approach

### First Aid course, optional for individuals who have recent first-aid training or corresponding professional background

- Theoretical and practical training in First Aid, following current guidelines from the Swedish Resuscitation Council (51), adhering to the First Aid guidelines of the European Resuscitation Council (52)

The whole course is held by experienced CPR and First Aid instructors certified by the Swedish Resuscitation Council. The study-specific course has been reviewed by the course instructors to improve quality of the course material and consistency in the assessment of eligible participants.

**Figure 2.**
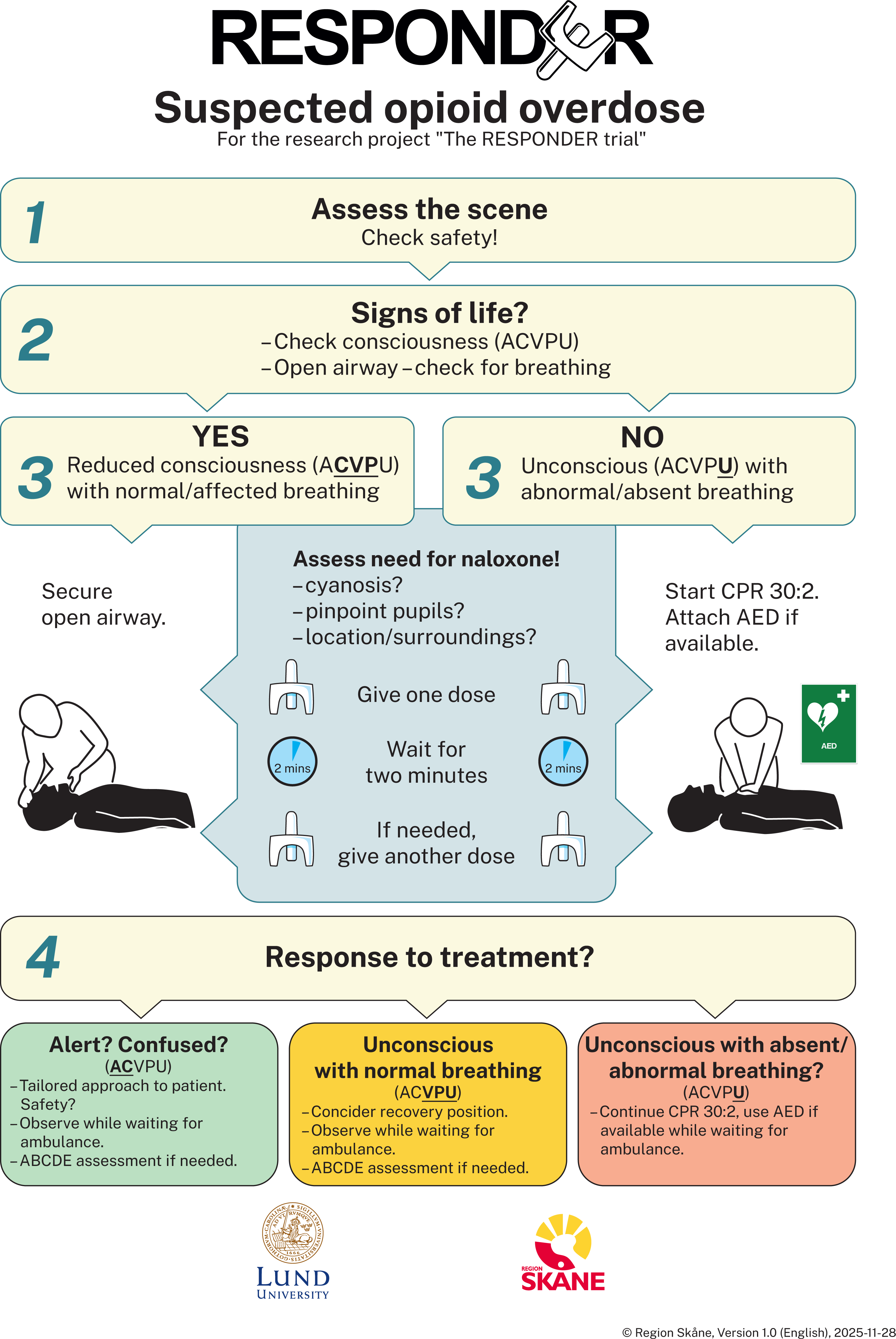
Study-specific naloxone algorithm

Eligible participants will be provided with a kit containing one pair of disposable gloves, a hands-on brochure with the naloxone algorithm, contact information to the study group, dry and wet wipes, and a pocket mask for manual ventilation. In line with current regulations, two doses of intranasal naloxone containing 1.26 mg naloxone each (currently brand name Respinal), will be purchased by study participants from local pharmacies and added to the naloxone kit.

### Eligibility criteria – community first responders

Potential CFRs, hereafter called naloxone-responders in the context of study-specific CFRs, need to fulfill six criteria to become registered in the project:

1. Provide informed written consent to participate in the study
2. Successfully complete the previously described study-specific course
3. Demonstrate understanding of the importance of a low-arousal approach
4. Download and register oneself as a user in the study-specific CFR application on a smartphone
5. Purchase naloxone from a pharmacy and receive reimbursement for the full cost of the purchase upon submission of the pharmacy receipt
6. Be 18 years of age or older and a resident in Region Skåne, Sweden

To successfully complete the pre-registration training (no. 2), participants must show comprehension in discussions concerning the importance of low-arousal approach when interacting with people in distress, such as when experiencing withdrawal after overdose reversal (no. 3). The learning of low-arousal approach is held in agreement with regional TERMA (ThERapeutic Meeting with Aggression) guidelines, derived from the Bergen-model (53). Study-specific instruction videos with simulated overdose and OHCA scenarios are included in the course to encourage ethical discussions about approach and assessment during alerts.

An inter-rater reliability survey will be performed by the instructors and controlled by the study group, to secure a concordant assessment of successful completion of the course. Furthermore, course participants whose eligibility is uncertain—for example, due to insufficient understanding of the low-arousal approach—will be discussed between the instructor and the study’s steering group to determine suitability for participation.

### Eligibility criteria – opioid overdose

Naloxone-responders can be notified of suspected overdoses and OHCAs between 7 a.m. and 11 p.m. These standards agree with present procedures for CFRs in Sweden. Participants are advised to carry the naloxone kit at all times. Participants can manually disable notifications Overdose events with victims ≥15 years old can activate the RESPONDER system, while the age limit of OHCA events is 8 years of age. No victim <15 years old can participate in follow-up interviews or in follow-up identified data collections, but non-identifying information from naloxone-responders and dispatcher records can be collected. Overdose victims will be contacted six hours up to seven days after the incident and receive information about RESPONDER, and be asked about approval and written consent to 1) collect data from the individual’s hospital records from the event and 2) to share their experience via interviews.

Ambulance dispatchers activate the RESPONDER dispatch of naloxone-responders when involvement of opioids is suspected in emergencies with altered consciousness. Naloxone-responders will also adhere to the CFR service for OHCAs in the same way as it is conducted today, such that included study participants will be available for dispatch to both suspected OHCAs and opioid overdose events. Participants who are already enrolled in the CFR service for OHCAs will have their current smartphone application inactivated and replaced by the new project-related application. No alert will be sent to naloxone-responders if dispatchers suspect homicide, suicidality, in other ways dangerous or violent situations, if the environment is perceived to be unsafe, or in case of traumatic cardiac arrests. Prior to project initiation, ambulance dispatchers at the regional EMDC in Region Skåne, responsible for the manual dispatch of naloxone-responders, completed an e-learning course covering the RESPONDER trial’s objectives and eligibility criteria for dispatching naloxone-responders. The dispatchers also engaged in in-person discussions with study representatives to ensure clarity and alignment regarding the trial procedures.

### Outcomes

Primary outcomes are (1) number and proportion of true opioid overdoses with naloxone-responders administering naloxone successfully prior to EMS arrival, (2) positioning of and acceptability of alerts among naloxone-responders in relation to overdose locations, (3) enrollment, demographics and characteristics of participants, and (4) safety for naloxone-responders and overdose survivors.

Secondary outcomes are (5) safety and acceptability of the intervention as experienced by naloxone-responders and overdose survivors, and (6) clinical outcome of overdose victims in events occurring before versus after onset of the trial.

### Sample size – naloxone-responders

Due to the nature of opioid overdose events, prior knowledge on total amount of overdoses and their localization does not allow for a precise calculation of a sufficient sample size. Based on experience from CFR models for OHCA, and previous naloxone programs for patients at risk of overdose in Sweden, in line with the best assumptions possible, a total of 500-1000 naloxone-responders will be included. Each naloxone-responder will be followed for at least six months and at most 12 months.

### Patient and Public Involvement Statement

Public communication about the trial will be displayed through communication in regional commuting areas, the website of Region Skåne, newspapers, radio, television reportages and social media platforms, aiming to reach and recruit a variety of individuals.

A reference group will follow the study and will be made up of representatives of several stakeholders in relation to the study topic, representatives of peer support groups in psychiatry generally and in opioid use disorders specifically, decision makers in psychiatry and addiction health care services, EMS professionals, and a peer support organisation in the area of cardiac and pulmonary diseases. In addition, political decision makers and administrative officers representing a wide range of health care administration in Region Skåne are thoroughly informed about the project continuously and will provide possibilities for a rapid implementation process in case of favorable study results.

### Data collection and management

Included data is shown in Table 1 and further explained below.

**Table 1.** Data inclusion table

|  |
| --- |
| **1. Naloxone-responder** |
| Post-alert questionnaire: use of naloxone, arrival time compared to emergency service, perception of situation and vital status of victim among other questions |
| Post-alert telephone call with RESPONDER team member |
| 12 months follow-up questionnaire |
| Qualitative interviews after participation |
| Age, sex, employment, prior medical and overdose-related experience, way of recruitment, contact details |
| **2. Individual affected by and surviving an opioid overdose, with naloxone-responder at the scene** |
| Post-alert contact with RESPONDER team member for asked participation in interview |
| **3. Pre-hospital records** |
| Time of arrival compared to responder |
| Age, sex and vital parameters (heart frequency and rhythm, respiratory frequency, blood pressure, level of consciousness, oxygen saturation) of overdose victim on site |
| Medications including naloxone |
| Transport or not to hospital |
| **4. Hospital records - With informed written consent from overdose survivor** |
| Clinical outcome, complications and survival until hospital discharge |
| Neurological function up to 48 hours after the event or until discharge from hospital |
| **5. Naloxone-responder application** |
| Total number of alerts |
| Number of accepted versus rejected alerts |
| Distance between naloxone-responder and alert |
| Location of naloxone-responder |
| Location of alert |
| **6. Emergency Medical Dispatch Centre data** |
| Dispatch code and possible key terms for suspected opioid overdose in call |
| Location of alert |
| Time between alert and emergency service arrival |

Two electronic surveys will be sent out to naloxone-responders 1.5 hours after every suspected overdose or OHCA alert, one of which is a routine survey already sent out to CFRs after OHCA events today, and one additional survey with questions related to suspected overdose events.

Included overdose-specific questions will address time of arrival (prior, simultaneously or after EMS), naloxone given or not (0, 1 or 2 doses), causes for not giving naloxone if so, impression of the medical situation (opioid overdose or other emergency), impression of clinical outcome, user experience, and safety at the scene. Experience and safety will include prevalence of reported adverse events in terms of threat, physical injury and long-lasting severe emotional distress. Naloxone-responders will also be dialed, after attending suspected emergencies, by a study group member with experience in mental health support. Open-ended questions regarding participants’ experiences, suggestions for improvement, and potential need for psychological support will be explored.

A follow-up questionnaire will be addressed to naloxone-responders 12 months post registration, regardless of whether they have been dispatched during the study period or not. Experience of participating, number of accepted events and frequency of naloxone administered during alerts will be examined. It will also include questions about experience of the pre-registration training and the individual’s perceived preparedness for alerts during the study period.

Qualitative interviews will be carried out with naloxone-responders and surviving overdose victims, resulting in two separate studies. Both studies will explore experiences from situations involving naloxone-responders, from the naloxone-responders’ and the surviving overdose victims’ perspectives, respectively. Data collection will continue until saturation in the interview material is reached. The overdose victims will be identified through pre-hospital sources, informed about the trial and asked about consent to participate.

Ambulance records in the documentation software PARATUS will be collected for vital status at arrival, age and sex of the victim, number of administered naloxone doses and by whom, and clinical status after possible administration. Received vital parameters, if available, include heart frequency and rhythm, respiratory frequency, oxygen saturation, blood pressure and level of consciousness. Pharmaceuticals other than naloxone administered by EMS and whether the person was transported to hospital, refused to be transported or was already deceased, will also be viewed through pre-hospital records. Hospital records will be studied for clinical outcome, complications and survival up to 48 hours after overdose reversal or until discharge from hospital if an individual was hospitalized. The individual results will be compared in relation to the time elapsed from the dispatcher’s receipt of the emergency call to the administration of naloxone.

Technical dispatcher data and naloxone-responder data will also be collected for overdose location, distance between naloxone-responder and overdose, naloxone-responders’ response to alerts, and if any naloxone-responder arrived before EMS. Data will also be collected from the smartphone application and questionnaires to study naloxone-responders’ age, sex, prior knowledge and experience of addiction treatment and overdoses, employment, way of recruitment, home and work location.

Suspected overdoses identified by ambulance dispatchers will be compared with on-scene assessments made by naloxone-responders and EMS, using data from dispatcher, EMS, and hospital records, as well as questionnaires completed by naloxone-responders.

### Statistical methods

No hard endpoint data in terms of power calculations will be performed in this pilot context. Descriptive statistics will primarily be used.

### Ethical considerations

The Swedish Ethical Review Authority has approved the trial (file number 2024-05887-01), and the presentation of the trial adheres to the SPIRIT guidelines (suppl. 1) (54).

Both naloxone-responders and overdose survivors will receive information about the trial, and potential benefits and risks of inclusion. Naloxone-responders need to consent to participate in the trial. Identifiable clinical outcome data of individuals affected by overdose will be included only if the patient can be recognized and provides informed written consent. Anonymized data will be used from pre-hospital records and SOS Alarm.

In accordance with the CFR model for OHCAs in Sweden, participants can be dispatched to children ≥8 years old with suspected OHCAs. The age limit for dispatch to suspected opioid overdoses, and for study inclusion as an overdose victim, will be ≥15 years old. A low to non-existing number of cases among children are estimated in terms of overdoses. However, harmful use of opioids occurs among young teenagers. The distributed naloxone compound is only formally indicated for administration to adults in Sweden (55), despite prior evidential studies on naloxone’s safety among pediatric populations (21, 22). The availability of naloxone over the counter, together with legal provisions, allows its administration to anyone in need.

Previous studies on CFRs for OHCA have reported mental distress among some CFRs following fatal OHCAs, and in situations when CFRs were unable to accept a dispatch. However, most people report that the benefits of being able to help override the potential psychological impact (56, 57). Contact information to the study group will be included in the post-survey form after every accepted alert, to make it easier for people to seek support. Nurses, physicians and counsellors from the study group will be able to support participants experiencing emotional distress. Further interventions to sufficiently support individuals will be of high priority if wished for or considered needed. Contact information to the study group will also be easily available on the trial website (www.skane.se/psykiatri/responder).

Address specifications including door code, floor number and other essential information can be sent to naloxone-responders when dispatched, to secure a time-effective arrival. The information will be removed from the application after the dispatch or if the naloxone-responder rejects the alert. Detailed personal information about the victim will not be sent. The eligibility of presented information aligns with routines from the CFR system for OHCA in Sweden.

## DISCUSSION

### Differences from ongoing and prior overdose dispatch studies

The one-day course of RESPONDER makes the intervention globally unique. An understanding of the low-arousal approach is expected to be an important factor when communicating with an overdose victim experiencing distressful withdrawal. The risk of recurrent overdose once the naloxone’s effects diminish highlights the need for a calm and informative approach by bystanders towards overdose victims. Previous efforts to improve naloxone accessibility have generally not included training in a low-arousal approach within overdose management programs, if any training was offered.

Around half of opioid overdoses are currently not reported to the EMS. The two largest THN programs in Sweden observed just below 50% of participating patients calling EMS when witnessing an overdose (6, 33). Achieving complete coverage of overdoses is unlikely. A study by McAuley et al. in Scotland, 2017, measured the number of ambulance attendances to opioid overdoses before and after implementation of a national THN program. No significant difference in frequency of emergency calls was observed (58).

The PulsePoint program in the United States added nasal naloxone kits to some public holders of AEDs (59). If the concept of naloxone-responders is feasible and with high acceptability and safety for both naloxone-responders and overdose victims, a future ambition could be to implement public naloxone boxes in the uptake area of the RESPONDER trial. This may improve accessibility for bystanders to intervene in the case of a nearby opioid overdose. We also suspect that not all naloxone-responders will consistently carry their naloxone kits with them. Public holders with naloxone may increase the odds of naloxone being available to dispatched naloxone-responders.

A recently published randomized controlled trial protocol describes recruitment of registered PulsePoint members to become carriers of naloxone (39). A limitation of this model is the risk of being dispatched to a false-positive overdose call, although the situation may still warrant First Aid due to other emergent needs. The accuracy of opioid overdose recognition among dispatchers is as of today an underexplored field. The pre-registration course in the RESPONDER trial may improve First Aid readiness during alerts. Dispatched naloxone-responders increase their chance to act purposefully in situations of false-positive opioid overdoses, while the naloxone and overdose in-depth course helps naloxone-responders to properly identify and manage true overdoses.

One current issue among laypeople with no prior overdose experience is understanding of a commonly used medical term describing pupil size during opioid overdoses. Common knowledge of this term, *pinpoint pupils (miosis)*, was examined through a field study. Randomly selected passersby outside of a common train station in Malmö were interviewed one day in May 2025 (Suppl. 2). One third out of fifty participants accurately defined the size of pinpoint pupils and chose the correct illustration out of three alternatives. However, 12% claimed knowledge of the definition, but chose an incorrect illustration of the condition. The findings resulted in clarified course material about pupil size in the RESPONDER model, to properly help potential naloxone-responders to successfully recognize alerts with true opioid overdoses and need of naloxone. First Aid knowledge and understanding of opioid overdose assessment and naloxone-induced withdrawal is something we consider vital for a possible successful, broad implementation of an overdose dispatch system.

The study-specific course is essential for the RESPONDER trial. A belief is that increased knowledge in opioid overdose assessment and management along with discussions on ethical and legal aspects may prepare participants for complex situations. The included First Aid course increases the number of people with potentially life-saving knowledge during emergencies where they may become bystanders. Most of today’s CFR systems include all people who report that they know CPR (41, 46, 60). The new model of RESPONDER guarantees that participants have received up to date First Aid and CPR knowledge. The one-day course offers in-depth knowledge that no other trial with CFRs carrying naloxone has tried (39, 61–63). However, the included one-day course requires both work-related and economical efforts, and a greater number of participants demands additional course sessions. These course-related aspects may cause limitations regarding recruitment and consequently feasibility of the model and will be explored during the trial.

Overdoses caused by high-potency opioids like fentanyl and nitazenes may require additional dosing of naloxone (21, 64, 65). A strength of this trial is thereby the co-working setting between volunteers and EMS, allowing quick assistance with further dosing by EMS. The median response time for EMS during OHCAs in Skåne in 2024 was 11 minutes (interquartile range 8-16) (20).

Other overdose dispatch systems have either targeted people who inject opioids or have required bystanders to be registered in the system to dispatch others (39, 62). One such is a community-based overdose dispatch system with registered CFRs equipped with naloxone in Philadelphia, United States (62). Registered users alerted other registrants through an application when witnessing an overdose, which reportedly increased the likelihood of early reversal while awaiting EMS. Out of 291 suspected opioid overdoses, 30.6% were false positives. Among true positive cases, 95.9% achieved successful reversal and other registered CFRs arrived at least five minutes prior to EMS in 59.5% of times. A limitation, however, was that only registered users could activate the system. The RESPONDER trial is unique through its integration with EMDC and may reach additional overdose events, since ambulance dispatchers activate the system.

## CONCLUSIONS

The RESPONDER trial is a novel method for early dispatch of CFRs to the scene of suspected opioid overdoses and OHCAs using a smartphone-based dispatch system. These trained CFRs are equipped with the efficient antidote naloxone and may reach affected individuals at an earlier stage before EMS arrival and with greater coverage to sufficiently reduce mortality and morbidity from overdoses. Based on experiences from smartphone alerting systems with committed CFRs, we believe that RESPONDER with its trained CFRs may reduce the critical time from opioid overdose to safe and efficient administration of naloxone.

## Abbreviations

AED: Automated external defibrillator, CFR: Community first responder, CPR: Cardiopulmonary resuscitation, EMDC: Emergency medical dispatch centre, EMS: Emergency medical services, OHCA: Out-of-hospital cardiac arrest, RESPONDER: REgion Skåne Preventing Overdose through Naloxone Distribution with Emergency Runners, THN: Take-Home Naloxone.

## Data Availability Statement

Data are available upon reasonable request.

## Author Contributions

CRediT: **Julia Rehn:** Conceptualization, Data curation, Investigation, Methodology, Project administration, Resources, Software, Validation, Visualization, Writing - original draft, Writing - editing – **Cecilia Andréll:** Conceptualization, Data curation, Funding acquisition, Investigation, Methodology, Project administration, Resources, Supervision, Visualization, Writing – review and editing **- Josephine Hansson:** Conceptualization, Methodology, Project administration,

Resources, Writing – review and editing **- Katja Troberg:** Conceptualization, Data curation, Investigation, Methodology, Project administration, Resources, Supervision, Validation, Visualization, Writing – review and editing **- Pernilla Isendahl:** Conceptualization, Resources, Writing – review and editing – **Martin Bråbäck:** Conceptualization, Supervision, Writing – review and editing **- Leif Svensson:** Conceptualization, Writing – review and editing **– Fredrik Byrsell:** Conceptualization, Methodology, Project administration, Resources, Writing – review and editing – **Andreas Claesson:** Conceptualization, Methodology, Writing – review and editing – **Anders Håkansson:** Conceptualization, Data curation, Funding acquisition, Investigation, Methodology, Project administration. Resources, Supervision, Writing – review and editing.

All authors have approved the final version of the manuscript.

## Acknowledgements

The authors wish to acknowledge Region Skåne, Practicum Clinical Skills Centre, and its instructors, for organizational support and expertise. The authors are grateful to Fredrik and Ingrid Thuring’s foundation for research funding.

The authors wish to thank SOS Alarm for the collaboration in implementing the model and integrating it with regular emergency medical dispatch service, and Heartrunner Sweden AB for collaboration regarding the smartphone application.

Also, thanks to Studio Korsholm, and Lund University academic training facility MedCUL (including medical licentiate Sten Erici), for their production of videos illustrating potential naloxone-responder scenarios. We also wish to acknowledge the whole video team including directors, actors and expertise from the clinical field. The authors are also grateful to communication staff or Region Skåne for their help with the graphic designs and information dissemination.

## Conflicts of Interest

This work was financially supported by grants from Region Skåne for AH, CA, KT and MB. The remaining authors had no financial support for this work. AH has overall study funding from the state-owned gambling operator of Sweden and from its research council, as well as from the research council of the Swedish alcohol monopoly, in projects entirely unrelated to the present study. AC and LS were shareholders in Heartrunner Sweden AB until July 2025. After this date, AC and LS have no commercial connections to the company. FB is an employee at the state-owned dispatching service SOS Alarm AB.

None of the remaining authors declare any conflict of interest regarding the current publication.

The study’s steering group and researchers will not be naloxone-responders themselves during the trial, due to potential participation bias.

## Notes

### Competing Interest Statement

This work was financially supported by grants from Region Skane for AH, CA, KT and MB. The remaining authors had no financial support for this work. AH has overall study funding from the state-owned gambling operator of Sweden and from its research council, as well as from the research council of the Swedish alcohol monopoly, in projects entirely unrelated to the present study. AC and LS were shareholders in Heartrunner Sweden AB until July 2025. After this date, AC and LS have no commercial connections to the company. FB is an employee at the state-owned dispatching service SOS Alarm AB.
None of the remaining authors declare any conflict of interest regarding the current publication.
The steering group and researchers will not be naloxone-responders themselves during the trial, due to potential participation bias.

### Clinical Trial

NCT07079241

### Funding Statement

This work was financially supported by grants from Region Skane for AH, CA, KT and MB. The remaining authors had no financial support for this work.

### Author Declarations

The Swedish Ethical Review Authority gave ethical approval for this work (file number 2024-05887-01).

